# Clinical Consequences of Occult Free Valproate Toxicity in Critically Ill Adult Patients: A Multicenter Retrospective Cohort Study

**DOI:** 10.1101/2024.05.16.24307506

**Authors:** Andrew J Webb, David J Gagnon, Caitlin S Brown, Richard R Riker, Natasha D Lopez, Melanie Z Goodberlet, Michael J Schontz, Kaylee K Marino, Sacha N Uljon, Sahar F. Zafar, Eric S Rosenthal

## Abstract

**Background and Objectives:** Valproate has wide pharmacokinetic variability and a narrow therapeutic index. Its protein binding is unpredictable, particularly among critically ill patients who may experience unexpectedly elevated free concentrations. We sought to identify the clinical consequences and determinants of occult free valproate toxicity in critically ill adults.

**Methods:** We conducted a multicenter retrospective cohort study of adult patients admitted to an intensive care unit (ICU) who were receiving valproate and had concurrent total and free valproate concentrations measured. We examined whether valproate concentrations were independently associated with adverse drug effects (ADEs) including thrombocytopenia, hepatotoxicity, hyperammonemia, and pancreatic injury. Determinants of occult toxicity were also identified using logistic mixed-effects models, adjusting for age, weight, albumin, propofol and aspirin use, and blood urea nitrogen (BUN). Occult toxicity was defined as a free valproate concentration that was discordant with total concentration (e.g., supratherapeutic free concentration associated with therapeutic total concentration).

**Results:** 311 unique patients (mean age 58 [±17] years, 36% female, 31% non-white, and 29% on valproate prior to admission) with 550 concurrent free and total valproate concentration pairs met inclusion criteria. The median (IQR) total valproate concentration was 46 mcg/mL (34-63) and the median free valproate concentration was 17 mcg/ml (11-23); median free fraction was 35% (25-63%). Eighty-four percent of total valproate concentrations represented occult free toxicity; a therapeutic total with a supratherapeutic free valproate concentration was the most common pattern (32% of concentration pairs). Each 2.5 mcg/mL increase in free valproate concentration was associated with thrombocytopenia (adjusted unit odds 1.15, 95% CI 1.05-1.26) and hepatotoxicity (adjusted unit odds 1.11, 95% CI 1.05-1.18). Albumin concentration (adjusted odds; aOR 0.17, 95% CI 0.08-0.36), BUN (aOR 1.36, 95% CI 1.09-1.70), and propofol exposure (aOR 3.06, 95% CI 1.38-6.79) were associated with occult toxicity.

**Conclusion:** Free valproate concentrations should be measured in critically ill patients because it is associated with ADEs and is often underrepresented by total concentrations. Most critically ill patients are at risk, especially those with hypoalbuminemia, uremia, and lipid exposure.

## INTRODUCTION

Valproate has been used to treat seizure, agitation, migraine, and bipolar disorder for over 45 years. Therapeutic drug monitoring is recommended to ensure serum concentrations are maintained within a narrow therapeutic window. The reference range of valproate (50-100 mcg/mL) reflects the “total” concentration of both protein-bound and unbound drug. Valproate has complex protein binding, and the total concentration may not accurately reflect the biologically active free concentration.^1,2^ Free valproate percent is approximately 5-15% in healthy outpatients but can exceed 50-70% in critically ill patients.^3–5^ High free valproate concentrations may lead to adverse drug effects (ADEs), especially when doses are titrated to the total reference range of 50-100 mcg/mL.^6–9^ Occult toxicity, in which the total concentration appears low or therapeutic but underestimates the free valproate concentration, occurs in up to 70% of critically ill patients, and may mislead providers to make dosing errors.^4^ Better characterization of the clinical consequences and determinants of occult toxicity may allow for the proper selection of patients to measure free valproate concentrations, especially in centers without in-house availability of the free valproate concentration assay. Accordingly, the objectives of this study were to assess the relationship between free valproate concentration and selected ADEs, and to identify determinants of occult free valproate toxicity in critically ill patients.

## METHODS

We conducted a retrospective cohort study of adult patients admitted to intensive care units (ICUs) at two academic medical centers in Boston, MA, USA. Patients were included if they were over the age of 18 years, admitted to an ICU (neurologic, medical, surgical, trauma, cardiac, cardiothoracic, or burn) between December 2015 and December 2023, received valproate in the ICU, and had total and free valproate concentrations measured concurrently. Patients were excluded if the free valproate concentration was unreportable (e.g., insufficient volume in the tube sent for assay). Only the first admission per patient was included if multiple admissions were identified.

### Standard Protocol Approvals, Registrations, and Patient Consents

The study was deemed exempt by the Massachusetts General Brigham Institutional Review Board and the need for informed consent was waived (2023P000131).

### Data Collection and Procedures

Baseline demographics, hospitalization characteristics, and all concurrently measured free and total valproate concentrations were collected. Past medical history was assessed using ICD-10 codes. Charlson comorbidity index scores were collected to provide a point estimate of premorbid health status and sequential organ failure assessment (SOFA) scores were collected at ICU admission to quantify severity of illness. Laboratory values collected included serum albumin, creatinine, and blood urea nitrogen (BUN), all assayed within 72 hours of valproate concentration measurement and all platelet count, transaminases, ammonia, and lipase values drawn during admission to assess for ADEs. Concomitant medications collected included aspirin, acetaminophen, phenytoin/fosphenytoin, carbapenem antibiotics, propofol (for any duration), clevidipine (for any duration), intravenous fat emulsion, ketorolac, or ibuprofen administered within 24 hours prior to valproate concentration measurement. Data were abstracted from the institution’s enterprise data warehouse and organized in a common REDCap instrument.^10,11^

Total valproate concentration measurements were performed in-house at both institutions but combined total and free concentration measurements (i.e., the measurements included in this study) were send-out assays to Mayo Clinic (Rochester, MN, USA) until August 5^th^, 2022, when Hospital 1 changed to utilize Quest Diagnostics (Marlborough, MA, USA). Both reference laboratories use a commercial homogenous enzyme immunoassay technique to measure total and free (after ultrafiltration) concentrations (Roche valproic reagent; Roche Diagnostics Corp, Indianapolis, IN). Neither institution had a protocol to routinely measure free valproate concentration at the time of the study.

### Study Objectives

The primary objective of this study was to determine if there was an association between total and free valproate concentrations and selected ADEs, including thrombocytopenia, hepatotoxicity, hyperammonemia, and pancreatic injury, in critically ill adults. Thrombocytopenia was defined as an absolute platelet count of less than 50,000 cells/µL, hepatotoxicity as aspartate aminotransferase (AST) or alanine aminotransferase (ALT) of greater than three times the upper limit of normal or alkaline phosphatase greater than two times the upper limit of normal, hyperammonemia as a plasma ammonia concentration of 60 µmol/L or greater, and pancreatic injury as a serum lipase of greater than three times the upper limit of normal. Normal reference ranges for laboratory values are summarized in **Table S1**. Any value collected within the seven days after the first free valproate measurement (and after initial valproate exposure) was eligible for assessment as a potential ADE and was compared against the highest measured free valproate concentration during that period. Patients who developed any of the ADEs prior to exposure to valproate were excluded from the regression analysis to limit confounding.

The secondary objective was to identify determinants of occult free valproate toxicity.

Occult toxicity was defined as a free valproate concentration therapeutic interpretation that was higher and discordant with its total concentration therapeutic interpretation (**Figure 1**). The reference range for total valproate concentration was 50-100 mcg/mL and for free valproate concentration was 5-15 mcg/mL. We utilized 5-15 mcg/mL as the reference range because it has been previously reported in similar studies and aligns with the expected free valproate concentration in the presence of normal protein binding.^4^ Determinants evaluated *a priori* included patient-specific factors (age, sex, race, weight, height, use of valproate prior to admission) and concentration-specific factors (albumin, BUN, and creatinine measurements within 72 hours of valproate concentration collection), and concomitant use of propofol, clevidipine, aspirin, NSAIDs, phenytoin, carbapenem antibiotics, or intravenous fat emulsion within 24 hours of valproate concentration collection.

**Figure 1:**
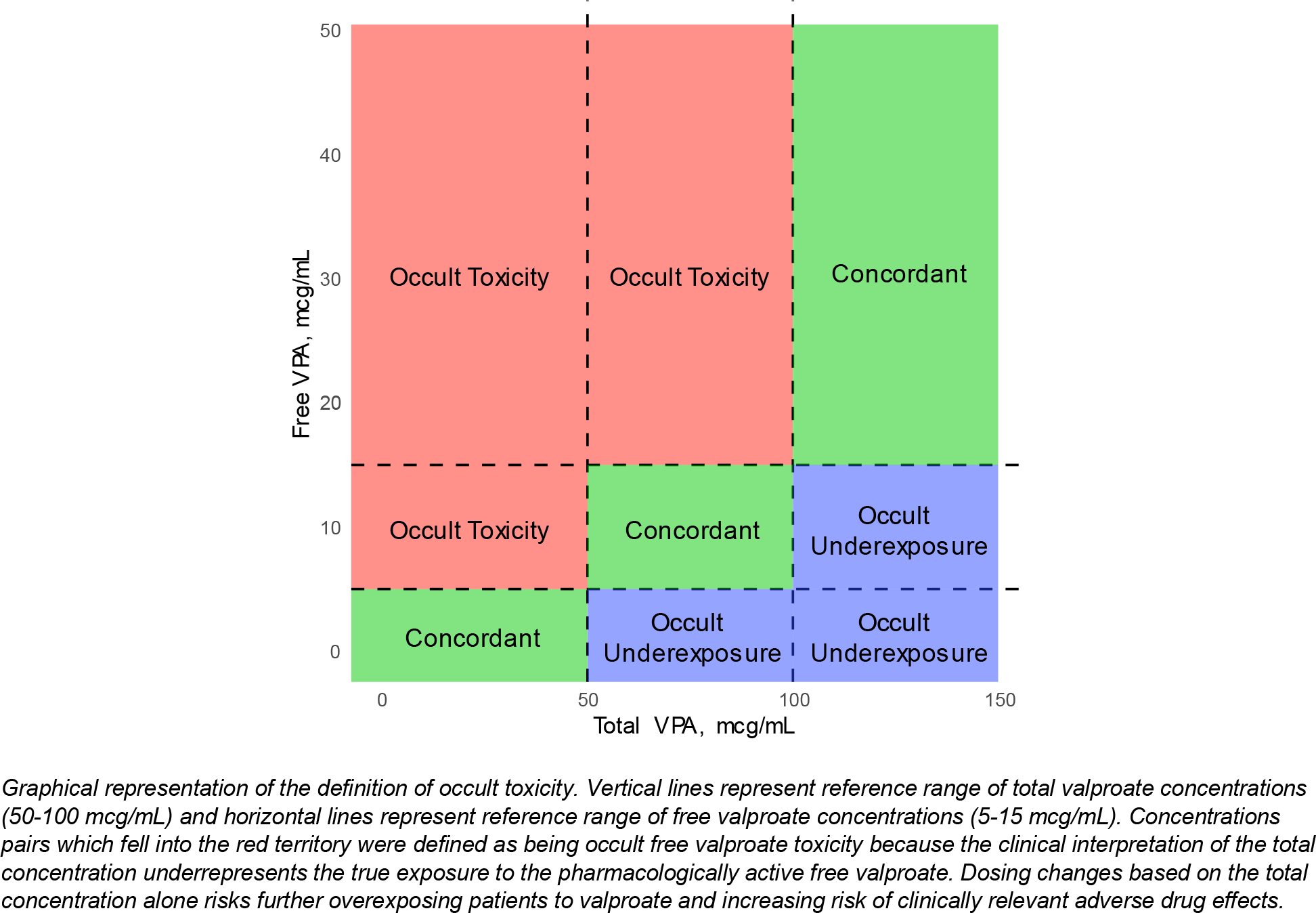
Outcomes Definition

### Statistical Analysis

Continuous data were summarized using means and standard deviations (SD) or medians and interquartile range (IQR) data, as appropriate. Nominal data were summarized using counts and percentages. P values less than 0.05 were statistically significant. A power calculation was not completed because the entire convenience sample of free and total valproate measurements was eligible for inclusion. The association between valproate concentrations and ADEs was assessed using multivariable logistic regression; only one concentration measurement per patient encounter (the highest within seven days of the first free valproate measurement) was included in the model. A sensitivity analysis was conducted excluding any patient who had the ADEs present at admission (regardless of whether the patient took valproate prior to admission) using just the first measurement for each patient. The association between valproate concentration (both total and free) and ADEs were adjusted for age, sex, and use of valproate prior to admission.

Independent determinants of occult toxicity were evaluated using logistic mixed effects models including relevant patient demographics and concentration-specific characteristics as fixed effects and individual patients as random effects. Factors with significant associations and those with a plausible biologic association with occult toxicity were carried into a multivariable logistic mixed effects model with patients as a random effect. Mixed effects models were constructed using the *lme4* package.^12^ Missing data were minimized as much as possible, and values were not imputed if missing and were considered missing at random. Only complete records were included in regression models. All analyses were conducted using R, version 4.1.2 (R Foundation for Statistical Computing, Vienna, Austria). Anonymized data not published within this article will be made available upon reasonable request from any qualified investigator after completion of a data use agreement.

## RESULTS

311 patients with 550 pairs of free and total valproate concentrations met analysis inclusion criteria. Six (2%) patients had multiple admissions during the study period, such that only the first admission was included for analysis. Baseline demographics are summarized in **Table 1**. Features of the laboratory measurements are summarized in **Table 2** and the relationship between measured total and free valproate concentrations is depicted in **Figure 2**. Of the 550 free-total valproate concentration pairs, the total and free concentrations were concordant in only 88 (16%) pairs. In 462 (84%) pairs, the total concentrations discordantly underestimated the free concentration. The total concentration never overestimated the free concentration category (e.g., total concentration therapeutic but free concentration subtherapeutic). A total valproate concentration within the reference range associated with a supratherapeutic free valproate concentration was the most frequent pattern of occult toxicity, occurring in 176 (32%) pairs.

**Figure 2:**
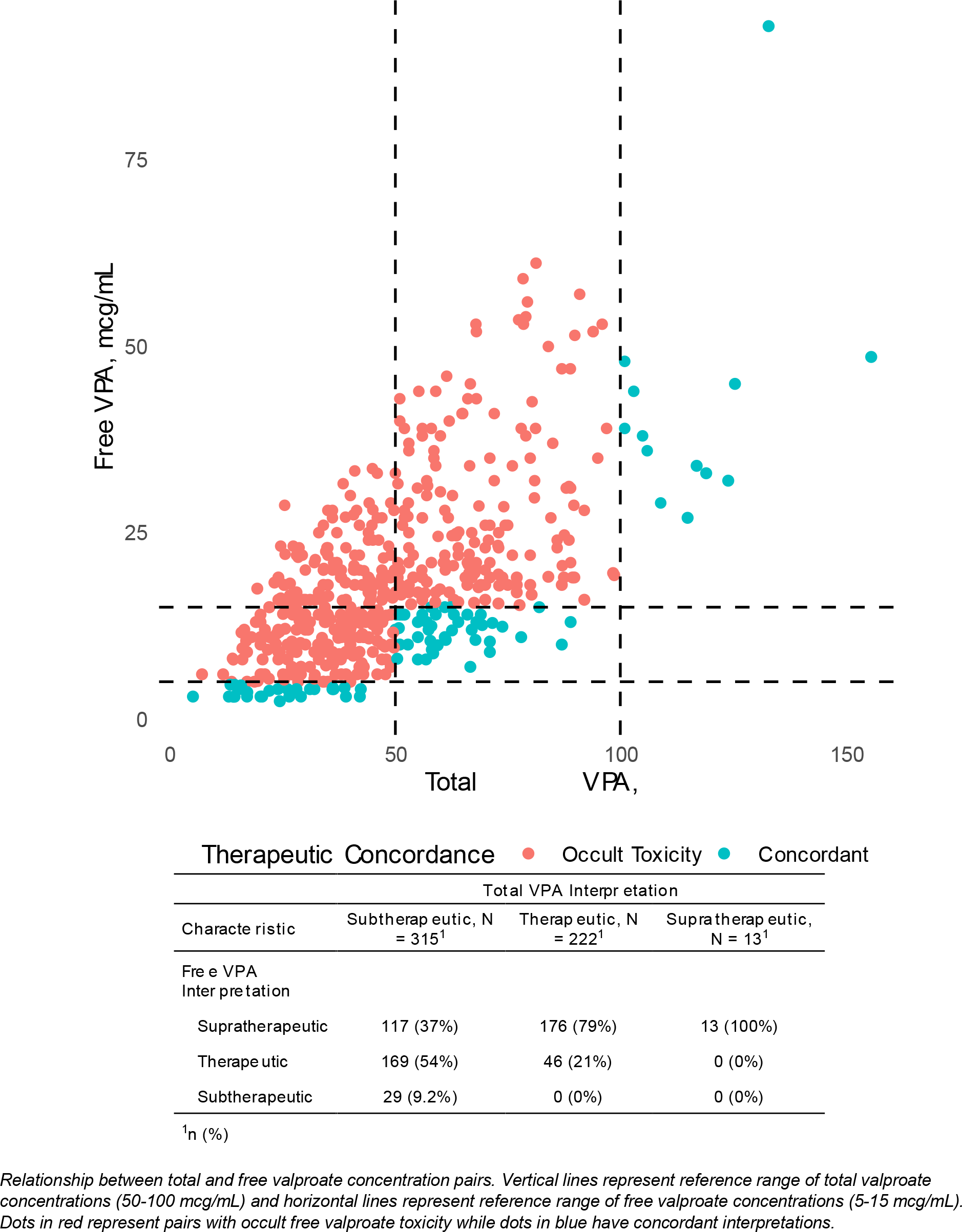
Total versus Free Concentrations

**Table 1:** Baseline Demographics.

| **Characteristic** | **Unique Admissions**<br>**N = 311<sup>1</sup>** |
| --- | --- |
| **Age, years** | 58 (17) |
| **Female, %** | 105 (34%) |
| **Race** |  |
| White | 216 (69%) |
| Black or African American | 33 (11%) |
| American Indian or Alaskan Native | 2 (0.6%) |
| Asian | 7 (2.3%) |
| Other/Unknown | 53 (17%) |
| **Weight, kg** | 81 (20) |
| **Site** |  |
| Hospital 1 | 217 (70%) |
| Hospital 2 | 94 (30%) |
| **On valproate Prior to Admission, %** | 90 (29%) |
| **Past Medical History** |  |
| Seizure Disorder | 103 (33%) |
| Psychiatric Disorder* | 65 (21%) |
| Heart Failure | 30 (9.6%) |
| Diabetes | 51 (16%) |
| CKD | 53 (17%) |
| Liver Disease^ | 17 (5.5%) |
| Alcohol use | 22 (7.1%) |
| **Admitting Service** |  |
| Neurology/Neurosurgery | 134 (43%) |
| Medical | 109 (35%) |
| Surgical | 47 (15%) |
| Cardiac/Cardiac Surgical | 19 (6.1%) |
| **Hospital Length of Stay, days** | 27 (15, 45) |
| **ICU Length of Stay, days** | 12 (6, 21) |
| **SOFA Score on Admission<sup>#</sup>** | 4 (1.8, 7.0) |
| **Charlson Comorbidity Index<sup>#</sup>** | 3 (1, 4) |
| **Discharge Disposition** |  |
| Home | 66 (21%) |
| Rehabilitation | 66 (21%) |
| LTC | 90 (29%) |
| Hospital Transfer | 4 (1.3%) |
| Hospice | 15 (4.8%) |
| Deceased | 70 (23%) |

<sup>1</sup>Mean (SD); n (%); Median (IQR)
\*Schizophrenia, bipolar disorder, or depression
^Cirrhosis, steatosis, or fibrosis
<sup>#</sup>Data unavailable for 95 patients
CKD: chronic kidney disease; ICU: intensive care unit; LTC: long term care facility; SOFA: sequential organ failure assessment

**Table 2:** Features of Free and Total Valproate Concentrations.

| Characteristic | Free/Total Valproate Interpretation* |  |  | p-value<sup>2</sup> |
| --- | --- | --- | --- | --- |
|  | Overall,<br>N = 550<sup>1</sup> | Occult Toxicity,<br>N = 462<sup>1</sup> | Concordant,<br>N = 88<sup>1</sup> |  |
| **Free valproate, mcg/mL** | 17 (11, 23) | 18 (12, 25) | 11 (4, 14) | <0.001 |
| **Total valproate, mcg/mL** | 46 (34, 63) | 44 (34, 61) | 57 (35, 69) | 0.011 |
| **Free fraction, %** | 35% (25, 63) | 39% (29, 55) | 20% (25, 26) | <0.00 |
| **valproate dose in Prior 24 Hours, mg** | 1,600 (1,000, 2,500) | 1,725 (1,000, 2,715) | 1,500 (1,000, 2,500) | 0.014 |
| **Albumin, g/dL<sup>^</sup>** | 2.80 (2.50, 3.10) | 2.70 (2.40, 3.00) | 3.10 (2.90, 3.60) | <0.001 |
| Missing | 45 | 41 | 4 |  |
| **BUN, mg/dL<sup>^</sup>** | 24 (16, 40) | 25 (16, 42) | 19 (12, 26) | <0.001 |
| Missing | 3 | 1 | 2 |  |
| **SCr, mg/dL<sup>^</sup>** | 0.81 (0.57, 1.47) | 0.85 (0.58, 1.83) | 0.69 (0.54, 0.90) | <0.001 |
| Missing | 3 | 1 | 2 |  |
| **Concomitant drugs** |  |  |  |  |
| Propofol | 159 (29%) | 147 (32%) | 12 (14%) | <0.001 |
| Phenytoin | 11 (2.0%) | 10 (2.2%) | 1 (1.1%) | >0.99 |
| Carbapenem | 17 (3.1%) | 12 (2.6%) | 5 (5.7%) | 0.17 |
| Clevidipine | 4 (0.7%) | 4 (0.9%) | 0 (0%) | >0.99 |
| Aspirin | 122 (22%) | 113 (24%) | 9 (10%) | 0.003 |
| Acetaminophen | 279 (51%) | 242 (52%) | 37 (42%) | 0.076 |
| Ibuprofen | 4 (0.7%) | 4 (0.9%) | 0 (0%) | >0.99 |
| Ketorolac | 1 (0.2%) | 1 (0.2%) | 0 (0%) | >0.99 |

<sup>1</sup>Median (IQR); n (%)
<sup>2</sup>Wilcoxon rank sum test; Pearson's Chi-squared test; Fisher's exact test
\*Concordance refers to the qualitative interpretation of the free and total level matching (e.g. both free and total level therapeutic). Free valproate therapeutic range defined as 5-15 mcg/mL, total valproate therapeutic range defined as 50-100 mcg/mL
<sup>^</sup>The closest lab value collected within 72 hours of free/total valproate level being drawn was included
BUN: blood urea nitrogen, SCr: serum creatinine

Of patients with evaluable laboratory data, 47/249 (19%) patients experienced hepatotoxicity, 10/308 (3.2%) experienced thrombocytopenia, 38/172 (22%) experienced hyperammonemia, and 7/83 (8.4%) experienced pancreatic injury (**Table S2**). Each 2.5 mcg/mL increase in free valproate concentration was associated with increased odds of thrombocytopenia (adjusted odds ratio; aOR 1.17, 95% CI 1.06-1.30) and hepatotoxicity (aOR 1.11, 95% CI 1.05-1.18) while free valproate concentrations were not associated with hyperammonemia or pancreatic injury (**Table 3**). Results were unchanged when patients with lab abnormalities present at admission were excluded from analysis (**Tables S3-4**). A negative correlation was observed between the free valproate concentrations and the lowest measured platelet count (**Figure S2**). Total valproate concentration (per 10 mcg/mL increase) was not associated with thrombocytopenia, hepatotoxicity, or pancreatic injury, but was associated with increased odds of hyperammonemia (aOR 1.21, 95% CI 1.04-1.41).

**Table 3:** Association of Free and Total Concentrations with Adverse Drug Effects.

| **Free valproate (Per 2.5 mcg/mL increase)** |  |  |  |
| --- | --- | --- | --- |
| **Adverse Drug Effect\*** | **aOR<sup>1</sup>** | **95% CI<sup>1</sup>** | **p-value** |
| **Thrombocytopenia** | 1.17 | 1.06, 1.30 | 0.003 |
| **Hepatotoxicity** | 1.11 | 1.05, 1.18 | <0.001 |
| **Hyperammonemia** | 1.00 | 0.92, 1.07 | >0.99 |
| **Pancreatic Injury** | 1.10 | 0.99, 1.25 | 0.09 |
| **Total valproate (Per 10 mcg/mL increase)** |  |  |  |
| **Adverse Drug Effect\*** | **aOR<sup>1</sup>** | **95% CI<sup>1</sup>** | **p-value** |
| **Thrombocytopenia** | 1.15 | 0.86, 1.49 | 0.3 |
| **Hepatotoxicity** | 1.00 | 0.87, 1.14 | 0.9 |
| **Hyperammonemia** | 1.21 | 1.04, 1.41 | 0.01 |
| **Pancreatic Injury** | 1.33 | 0.96, 1.90 | 0.09 |

<sup>1</sup>aOR = adjusted Odds Ratio, CI = Confidence Interval
ORs >1.0 indicate positive association with adverse effect; each model adjusted for age, sex, and pre-admission valproate use
\*Thrombocytopenia: platelet count < 50,000 cells/μL, hepatotoxicity: AST >120 (male) or >96 (female) U/L, ALT > 165 (men) or >99 (female) U/L, or alkaline phosphatase >230 (male) or >200 (female) U/L, hyperammonemia: plasma ammonia > 60 umol/L, pancreatic injury: lipase > 180 U/L
Patients who experienced any of the adverse events prior to exposure to valproate were excluded from the model

Total valproate concentrations were similar in the group with and without ADEs and a total concentration within the reference range (rather than above it) was not associated with the avoidance of ADEs. Only 2 (4.3%) of the 47 patients who developed hepatoxicity, 1 (10%) of the 10 patients who developed thrombocytopenia, 4 (10.6%) of the 38 patients who developed hyperammonemia, and 1 (14.3%) of the 7 patients who developed pancreatic injury had a supratherapeutic total valproate concentration (**Table S5**). Free valproate concentrations were higher and more likely to be supratherapeutic in patients who experienced ADEs compared to those without ADEs. Supratherapeutic free concentrations were seen in 34 (72%) of the 47 patients who developed hepatotoxicity, 8 (80%) of the 10 patients who developed thrombocytopenia, 29 (76%) of the patients who developed hyperammonemia, and 6 (86%) of the 7 patients who developed pancreatic injury (**Table S5**).

Total valproate concentrations, sex, albumin concentration, propofol use, aspirin use, and BUN were assessed as determinants of occult toxicity (**Table 4**). Albumin per 1 mg/dL decrease (aOR 0.17, 95% CI 0.08-0.36), propofol exposure within 24 hours of concentration measurement (aOR 3.06, 95% CI 1.38-6.79), and BUN per 10 mg/dL increase (aOR 1.36, 95% CI 1.09-1.70) were independently associated with occult toxicity. Total valproate concentration (per 10 mcg/mL increase) was not significantly associated with the odds of occult toxicity. An albumin less than 2.9 g/dL was associated with a 58.9% sensitivity and 77.3% specificity for occult toxicity. Albumin concentrations below 2.9 g/dL had a 92.8% positive predictive value for occult toxicity and concentrations above 2.9 g/dL had a 27.3% negative predictive value for occult toxicity (**Figure S3**).

**Table 4:**
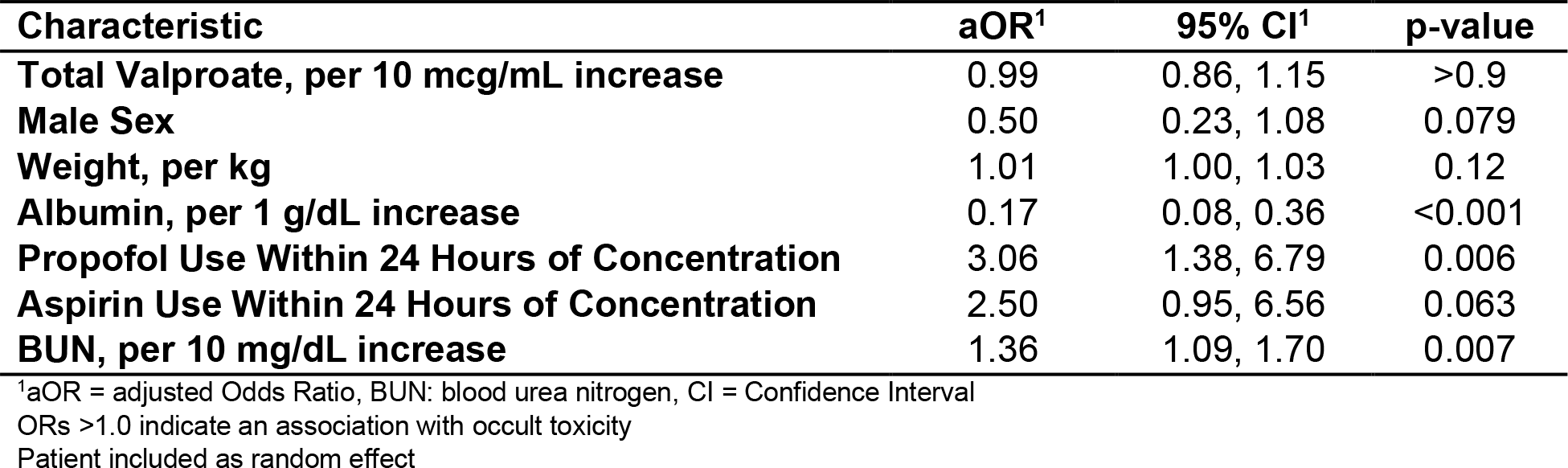
Determinants of Occult Toxicity.

| **Characteristic** | **aOR<sup>1</sup>** | **95% CI<sup>1</sup>** | **p-value** |
| --- | --- | --- | --- |
| **Total Valproate, per 10 mcg/mL increase** | 0.99 | 0.86, 1.15 | >0.9 |
| **Male Sex** | 0.50 | 0.23, 1.08 | 0.079 |
| **Weight, per kg** | 1.01 | 1.00, 1.03 | 0.12 |
| **Albumin, per 1 g/dL increase** | 0.17 | 0.08, 0.36 | <0.001 |
| **Propofol Use Within 24 Hours of Concentration** | 3.06 | 1.38, 6.79 | 0.006 |
| **Aspirin Use Within 24 Hours of Concentration** | 2.50 | 0.95, 6.56 | 0.063 |
| **BUN, per 10 mg/dL increase** | 1.36 | 1.09, 1.70 | 0.007 |

<sup>1</sup>aOR = adjusted Odds Ratio, BUN: blood urea nitrogen, CI = Confidence Interval
ORs >1.0 indicate an association with occult toxicity
Patient included as random effect

The relationship between propofol exposure, albumin, and valproate free fraction was further explored and is depicted in **Figure 3**. In patients without propofol exposure, valproate free fraction had a negative, linear correlation with serum albumin. Increasing doses of propofol appeared to magnify this effect, with valproate free fraction consistently being higher in patients receiving increasing doses of propofol.

**Figure 3:**
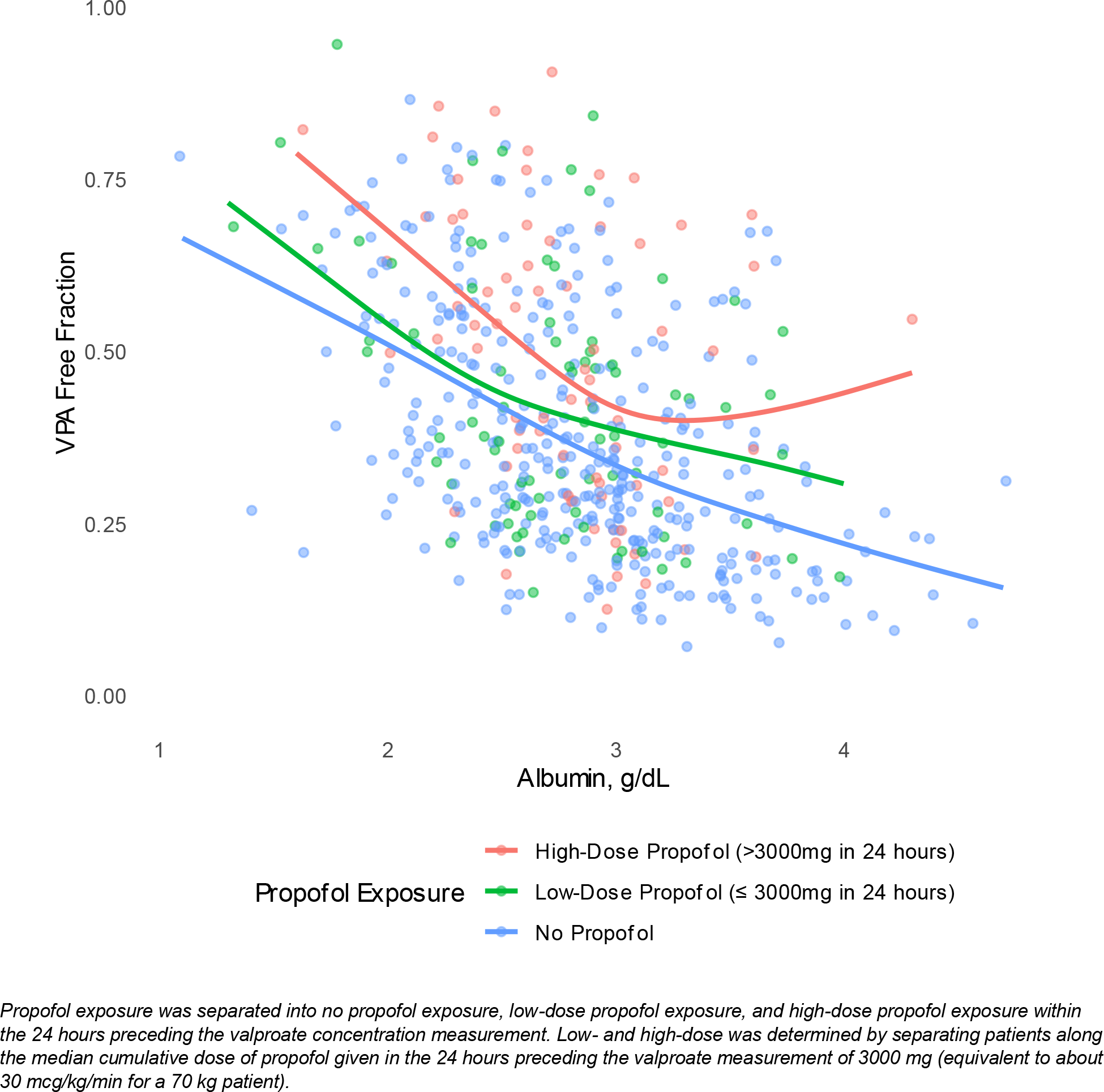
Effect of Albumin and Propofol Dose on Valproate Free Fraction

The observation that sex may contribute to the risk of occult toxicity (aOR 0.50, 95% CI 0.23-1.08 for male sex, **Table 4**) was also investigated (**Table S7**). Female patients received smaller median valproate doses in the 24 hours preceding concentration measurement (1500 vs 1750 mg, p=0.02) which may have been explained by lower body weight (68 vs 83 kg, p<0.01). Female patients had similar median total valproate concentrations compared to male patients (47 vs 45 mcg/mL, p=0.35) but both median free valproate concentration (20 vs 16 mcg/mL, p<0.002) and free fraction (40% vs 33, p=0.004) were significantly higher in female patients.

This was not explained by differences in serum albumin (2.80 vs 2.80 g/dL, p=0.75) or propofol (34% vs 27%, p=0.073) or aspirin (22% vs 22%, p=0.68) exposure between female and male patients, respectively, and BUN (20 vs 25 mg/dL, p<0.001) was lower in female patients.

## DISCUSSION

In this retrospective multicenter analysis of 311 critically ill patients, total valproate concentrations systematically underrepresent exposure to the pharmacologically active free valproate concentration; of 550 pairs of total and free valproate concentrations, 84% represented occult free valproate toxicity. Increasing free valproate concentrations were associated with the development of thrombocytopenia and hepatotoxicity while total valproate concentration was not associated with these ADEs. After adjusting for total valproate concentration, sex, and weight, the determinants independently associated with occult toxicity included decreased serum albumin, propofol exposure in the preceding 24 hours, and increasing BUN. These data suggest that using total valproate concentrations to guide dosing for critically ill patients may increase the risk of overdosing and increase the risk of ADEs.

The high rate of occult free valproate toxicity in our cohort demonstrates the complexity of valproate’s pharmacokinetics, especially in critically ill patients. Valproate protein binding is particularly variable; the free fraction in healthy patients is reported to range from 5 to 15%.^3^ In the ICU setting, however, many factors affect protein binding including hypoalbuminemia, uremia, or the administration of interacting medications such as aspirin and propofol or other lipid-containing medications. These factors reduce available albumin protein binding sites, resulting in an increased free valproate fraction which would not be identified if only a total valproate concentration is monitored. The median free fraction in our cohort was 35%, two- to seven-fold higher than the expected value in healthy adults. This is consistent with other critically ill cohorts, such as those reported by Gibbs (28.8%),^13^ Riker (48%),^5^ and Brown (23.6%).^4^

Rates of occult toxicity in hospitalized patients vary from 29.7% to 87%; the high rate of 84% occult toxicity in our population may reflect a higher severity of illness, as many prior cohorts reported a mixed population of non-ICU and critically ill adults.^4,5,13–16^ Additionally, our cohort had an overall low serum albumin (median 2.8 g/dL) and a high use of interacting medications (29% with propofol and 22% with aspirin exposure). This may reflect a selection bias in which free valproate concentrations were more likely to be measured in patients with these determinants, but our pattern of occult toxicities is consistent with previous reports, further enforcing our findings.^4,13^ This suggests that titrating valproate doses to a target total reference range of 50-100 mcg/mL may overexpose critically ill patients to free valproate, potentially increasing their risk for ADEs.

Higher free valproate concentrations were associated with ADEs in our cohort. Each 2.5 mcg/mL increase in free valproate concentration was associated with a 17% increase in the odds of thrombocytopenia and a 11% increase in the odds of hepatotoxicity. The association with hepatotoxicity was also independent of albumin concentration when this was added to the model. This differs from a prior study where no significant association between free valproate concentrations and ADEs was identified, but the smaller sample size (n=221) and low event rate in that cohort may explain the differing observations.^4^ We also used a more stringent definition of thrombocytopenia (<50,000 vs. <140,000 cells/µL) which improved our ability to discern clinically relevant ADEs. When we used a less strict platelet threshold was used to define thrombocytopenia (<140,000), the effect remained consistent in our data (aOR 1.20, 95% CI 1.09-1.39). Our findings are consistent with a prior publication that revealed a negative linear relationship between free valproate concentration and platelet count in outpatients.^17^

Total valproate concentrations were not associated with thrombocytopenia or hepatotoxicity and avoidance of supratherapeutic total valproate concentrations did not protect patients from developing these ADEs. However, total valproate concentrations were associated with hyperammonemia (aOR 1.21, 95% CI 1.04-1.41) while free concentrations were not. The mechanism for this discrepancy is unclear, as is the underlying mechanism of valproate-induced hyperammonemia.^18^ Valproate metabolites, including propionate, 4-en-valproate, and valproyl-CoA, may decrease hepatic N-acetylglutamate levels, subsequently impairing the mitochondrial urea cycle. Higher total valproate and 4-en-valproate concentrations were correlated with increasing plasma ammonia concentrations in one study of 11 pediatric patients with epilepsy but we did not replicate that finding or obtain 4-en-valproate concentrations in our study.^19^ Higher total valproate concentrations could reflect a higher proportion of metabolites not reflected in the higher free valproate concentrations, although we cannot confirm this theory. It is also possible that hyperammonemia is an idiosyncratic ADE with no correlation with either total or free valproate serum concentrations and our finding is due to chance.

Our findings suggest that free valproate concentrations should be directly measured in critically ill patients. Identification of factors associated with occult toxicity can improve appropriate patient selection if universal free valproate measurement is not feasible. Serum albumin was the strongest determinant of occult toxicity; the odds of occult toxicity increased by 83% for each 1 g/dL decrement in albumin. This is consistent with early reports of the importance of albumin for valproate protein binding; albumin is the principal protein responsible for valproate protein binding. Klotz and colleagues reported a free fraction of 29% in patients with alcohol-related liver disease and hypoalbuminemia which was significantly higher than the free fraction of 11% observed in healthy controls.^20^ Serum BUN, a surrogate for uremic toxins that compete for albumin binding sites,^21^ was also found to be associated with a 36% increase in the odds of occult toxicity for each 10 mg/dL increase. Both serum creatinine and BUN have been associated with increased free valproate concentrations, but BUN provides a closer representation of the biologic cause of binding displacement.^3,22^

Propofol exposure was associated with an approximately 3-fold increase in the odds of occult toxicity, although the confidence interval of this effect size was larger than the other variables tested (95% CI 1.38-6.79). Propofol, which is solubilized in 10% soybean oil, increases concentrations of free fatty acids which compete for valproate protein binding sites on albumin.^23,24^ We identified a significant association between propofol and free valproate fraction. Brown and colleagues revealed propofol exposure was associated with an increase in absolute free valproate concentration of 2.14 mcg/mL, but this was not significant (95% CI −0.99, 5.27), likely due to their smaller cohort of patients exposed to propofol.^4^ In an exploratory analysis of the effect of exposure level (**Figure 3**), higher exposure to propofol had a larger impact on free fraction, supporting this theory. Aspirin also demonstrated a trend towards occult toxicity in our cohort, but the effect did not reach statistical significance (aOR 2.50, 95% CI 0.95-6.56).

Aspirin’s lack of significant effect on occult toxicity may relate to most patients (66%) receiving low-dose (81 mg) aspirin; only 28.8% received 325 mg daily, and anti-pyretic dosing was uncommon (1.6%). While low-dose aspirin has been reported to lead to clinically meaningful valproate toxicity from increases in free fraction, this is limited to case reports and most higher doses of aspirin are generally required to influence valproate protein binding.^22,25–28^

We observed a significantly higher free fraction of valproate in female patients (40% vs 33%, p = 0.004) and a signal that male sex could be protective against occult toxicity, although this did not reach significance (aOR 0.50, 95% CI 0.23-1.08). Interestingly, absolute rates of occult toxicity were similar between sexes (87% vs 83%, p=0.29) despite the higher valproate free fraction. To our knowledge, this is the first description of an association between sex and valproate protein binding. Sex-based differences in protein binding are uncommon but may be related to changes in albumin or alpha-1-glycoprotein influenced by testosterone and estrogen.^29^ It is not clear if this difference is clinically significant but it should be explored in future studies.

Our study has several limitations. First, the decision to measure free and total valproate concentrations rather than a total concentration alone may be affected by referral or selection bias and the rate of occult toxicity may be inflated. However, the rate of occult toxicity reported here is within the reported range in other studies, including studies where a more universal free valproate testing strategy is employed,^4^ thus our findings are likely externally valid. There is also no universally accepted reference range for free valproate concentrations. While the reference range of 50-100 mcg/mL for total valproate is commonly used, reported free valproate reference ranges vary, including 4.3-17.3 mcg/mL,^13^ 5-17 mcg/mL,^5^ 5-10 mcg/mL,^14^ 4-12 mcg/mL,^15^ 7-23 mcg/mL,^16^ and 5-15 mcg/mL.^4^ Although an incorrectly low free valproate reference range might explain the high rate of occult toxicity we observed, the strong association between supratherapeutic free concentrations and ADEs suggests the range we selected should not be raised higher. An established reference range needs to be validated in prospective studies. The total reference range of 50-100 mcg/mL was also ascertained through conjecture from a sample of 13 outpatients with epilepsy in 1975,^30^ and the appropriateness of this range should be reassessed in this population. Lastly, clinical effectiveness (seizure cessation, agitation control) could not be reliably ascertained retrospectively, and thus the relationship between free valproate concentrations and clinical effect could not be determined. Given the complexity of these endpoints, this should be evaluated in a prospective study.

## Conclusion

Occult free valproate toxicity is common in critically ill patients. Dose titration to target a total concentration of 50-100 mcg/mL may overexpose critically ill patients to valproate. Patients with low albumin, renal dysfunction, and exposure to lipid-containing solutions such as propofol may benefit from direct measurement of free valproate concentrations, and avoidance of high free valproate concentrations may reduce the likelihood of adverse drug effects. The utility of routine free valproate concentration monitoring needs to be validated in a prospective study.

## Data Availability

Anonymized data not published within this article will be made available upon reasonable request from any qualified investigator after completion of a data use agreement.

