## Supplemental Appendix for "Clinical Consequences of Occult Free Valproate Toxicity in Critically Ill Adult Patients: A Multicenter Retrospective Cohort Study"

### Supplementary Appendix

#### Table S1: Normal Reference Ranges for Key Laboratory Values

| Laboratory Value | Reference Range | |
|---|---|---|
| | **Male** | **Female** |
| **Aspartate Aminotransferase (AST)** | 10-40 U/L | 9-32 U/L |
| **Alanine Aminotransferase (ALT)** | 10-55 U/L | 7-33 U/L |
| **Alkaline Phosphatase** | 45-115 U/L | 30-100 U/L |
| **Ammonia** | 12-48 µmol/L | |
| **Platelet Count** | 150,000-400,000 cells/µL | |
| **Lipase** | 13-60 U/L | |

#### Table S2: Incidence of Adverse Reactions by Therapeutic Discordance Status

| Characteristic | **Overall**, N = 311[1] | **Occult Toxicity**, N = 256[1] | **Concordant**, N = 55[1] | p-value[2] |
|---|---|---|---|---|
| **Hepatotoxicity** | 47 (19%) | 40 (20%) | 7 (15%) | 0.48 |
| Unable to assess | 62 | 53 | 9 | |
| **Thrombocytopenia** | 10 (3.2%) | 9 (3.6%) | 1 (1.8%) | >0.99 |
| Unable to assess | 3 | 3 | 0 | |
| **Hyperammonemia** | 38 (22%) | 29 (21%) | 9 (27%) | 0.42 |
| Unable to assess | 139 | 117 | 22 | |
| **Pancreatic Injury** | 7 (8.4%) | 6 (8.5%) | 1 (8.3%) | >0.99 |
| Unable to assess | 228 | 185 | 43 | |

[1]n (%)

[2]Pearson's Chi-squared test; Fisher's exact test

#### Table S3: Association of *Free* Valproate (per 2.5 mcg/mL increase) Concentration with Key Adverse Effects (Excluding Patients with Lab Abnormality Present on Admission)

| Adverse Effect* | aOR[1] | 95% CI[1] | p-value |
|---|---|---|---|
| **Thrombocytopenia** | 1.16 | 1.06, 1.29 | 0.002 |
| **Hepatotoxicity** | 1.11 | 1.05, 1.18 | <0.001 |
| **Hyperammonemia** | 1.02 | 0.96, 1.01 | 0.6 |
| **Pancreatic Injury** | 1.08 | 0.96, 1.22 | 0.2 |

[1]aOR = adjusted Odds Ratio, CI = Confidence Interval

ORs >1.0 indicate positive association with adverse effect; each model adjusted for age, sex, and pre-admission valproate use

*Thrombocytopenia: platelet count < 50,000 cells/µL, hepatotoxicity: AST >120 (male) or >96 (female) U/L, ALT > 165 (men) or >99 (female) U/L, or alkaline phosphatase >230 (male) or >200 (female) U/L, hyperammonemia: plasma ammonia > 60 umol/L, pancreatic injury: lipase > 180 U/L

**Table S4: Association of *Total* valproate (per 10 mcg/mL increase) Concentration with Key Adverse Effects (Excluding Patients with Lab Abnormality Present on Admission)**

| Adverse Effect* | aOR[1] | 95% CI[1] | p-value |
|---|---|---|---|
| **Thrombocytopenia** | 1.31 | 0.95, 1.77 | 0.081 |
| **Hepatotoxicity** | 1.00 | 0.87, 1.14 | 0.9 |
| **Hyperammonemia** | 1.20 | 1.00, 1.44 | 0.052 |
| **Pancreatic Injury** | 1.21 | 0.85, 1.68 | 0.2 |

[1]aOR = adjusted Odds Ratio, CI = Confidence Interval

ORs >1.0 indicate positive association with adverse effect; each model adjusted for age, sex, and pre-admission valproate use

*Thrombocytopenia: platelet count < 50,000 cells/µL, hepatotoxicity: AST >120 (male) or >96 (female) U/L, ALT > 165 (men) or >99 (female) U/L, or alkaline phosphatase >230 (male) or >200 (female) U/L, hyperammonemia: plasma ammonia > 60 umol/L, pancreatic injury: lipase > 180 U/L

### Table S5: Patterns of Total and Free Valproate Interpretation by Presence of Adverse Drug Effect (ADE)

| Characteristic | Hepatotoxicity | | p-value |
|---|---|---|---|
| | **ADE Not Present,** N = 202[1] | **ADE Present** N = 47[1] | |
| **Total Valproate Interpretation** | | | |
| Subtherapeutic | 106 (52%) | 26 (55%) | 0.76 |
| Therapeutic | 87 (43%) | 19 (40%) | 0.93 |
| Supratherapeutic | 9 (4.5%) | 2 (4.3%) | 0.47 |
| **Total Valproate Concentration** | 48 (35, 66) | 49 (33, 65) | 0.97 |
| **Free Valproate Interpretation** | | | |
| Subtherapeutic | 12 (5.9%) | 3 (6.4%) | 0.74 |
| Therapeutic | 75 (37%) | 10 (21%) | 0.07 |
| Supratherapeutic | 115 (57%) | 34 (72%) | 0.04 |
| **Free Valproate Concentration** | 18 (10, 25) | 22 (15, 34) | 0.017 |
| | **Thrombocytopenia** | | |
| | **ADE Not Present,** N = 298[1] | **ADE Present,** N = 10[1] | |
| **Total Valproate Interpretation** | | | |
| Subtherapeutic | 158 (53%) | 4 (40%) | 0.52 |
| Therapeutic | 129 (43%) | 5 (50%) | 0.75 |
| Supratherapeutic | 11 (3.7%) | 1 (10%) | 0.33 |
| **Total Valproate Concentration** | 48 (35, 65) | 52 (42, 65) | 0.48 |
| **Free Valproate Interpretation** | | | |
| Subtherapeutic | 16 (5.4%) | 0 (0%) | 0.99 |
| Therapeutic | 108 (36%) | 2 (20%) | 0.50 |
| Supratherapeutic | 174 (58%) | 8 (80%) | 0.20 |
| **Free Valproate Concentration** | 19 (11, 27) | 31 (20, 38) | 0.031 |
| | **Hyperammonemia** | | |
| | **ADE Not Present,** N = 134[1] | **ADE Present,** N = 38[1] | |
| **Total Valproate Interpretation** | | | |
| Subtherapeutic | 67 (50%) | 16 (42%) | 0.38 |
| Therapeutic | 62 (46%) | 18 (47%) | 0.90 |
| Supratherapeutic | 5 (3.7%) | 4 (11%) | 0.11 |
| **Total Valproate Concentration** | 50 (35, 63) | 59 (44, 75) | 0.012 |
| **Free Valproate Interpretation** | | | |
| Subtherapeutic | 8 (6.0%) | 0 (0%) | 0.20 |
| Therapeutic | 47 (35%) | 9 (24%) | 0.18 |
| Supratherapeutic | 79 (59%) | 29 (76%) | 0.051 |
| **Free Valproate Concentration** | 20 (11, 31) | 20 (16, 26) | 0.86 |
| | **Pancreatic Injury** | | |
| | **ADE Not Present,** N = 76[1] | **ADE Present,** N = 7[1] | |
| **Total Valproate Interpretation** | | | |
| Subtherapeutic | 42 (55%) | 4 (57%) | 0.99 |
| Therapeutic | 32 (42%) | 2 (29%) | 0.69 |
| Supratherapeutic | 2 (2.6%) | 1 (14%) | 0.23 |
| **Total Valproate Concentration** | 48 (28, 61) | 47 (44, 75) | 0.25 |
| **Free Valproate Interpretation** | | | |
| Subtherapeutic | 4 (5.3%) | 0 (0%) | 0.99 |
| Therapeutic | 29 (38%) | 1 (14%) | 0.41 |
| Supratherapeutic | 43 (57%) | 6 (86%) | 0.23 |
| **Free Valproate Concentration** | 20 (10, 28) | 27 (19, 42) | 0.19 |

[1]n (%); Median (IQR), [2]Fisher's exact test; Chi square test; Wilcoxon rank sum test

**Thrombocytopenia: platelet count < 50,000 cells/µL, hepatotoxicity: AST >120 (male) or >96 (female) U/L, ALT > 165 (men) or >99 (female) U/L, or alkaline phosphatase >230 (male) or >200 (female) U/L, hyperammonemia: plasma ammonia > 60 umol/L, pancreatic injury: lipase > 180 U/L*

**Table S6: Univariable Logistic Regression for Determinants of Discordance**

| Characteristic | N | OR[1] | 95% CI[1] | p-value |
|---|---|---|---|---|
| **Total Valproate, per 1 mcg/mL** | 550 | 0.84 | 0.76, 0.92 | <0.001 |
| **Male Sex** | 550 | 0.75 | 0.43, 1.26 | 0.3 |
| **Albumin, per 1 g/dL** | 505 | 0.21 | 0.13, 0.33 | <0.001 |
| **BUN, per mg/dL** | 547 | 1.04 | 1.02, 1.06 | <0.001 |
| **Weight, per kg** | 550 | 1.16 | 1.04, 1.31 | 0.011 |
| **Age, per year** | 550 | 1.01 | 1.00, 1.03 | 0.10 |
| **Medications Administered within 24 Hours of Concentration** | | | | |
| Propofol | 550 | 2.96 | 1.62, 5.87 | <0.001 |
| Aspirin | 550 | 2.84 | 1.45, 6.25 | 0.005 |
| Acetaminophen | 550 | 1.52 | 0.96, 2.42 | 0.077 |
| Phenytoin | 550 | 1.92 | 0.36, 35.6 | 0.5 |
| Carbapenem | 550 | 0.44 | 0.16, 1.42 | 0.14 |
| Ibuprofen | 550 | NC | 0.00, NA | >0.9 |
| Ketorolac | 550 | NC | 0.00, NA | >0.9 |

[1]OR = Odds Ratio, CI = Confidence Interval, BUN = blood urea nitrogen

NC: Not calculable

**Table S7: Association Between Sex and Valproate Concentrations**

| Characteristic | **Overall, N = 550[1]** | **Male, N = 393[1]** | **Female, N = 157[1]** | **p-value[2]** |
|---|---|---|---|---|
| **Free Valproate, mg/L** | 17 (11, 23) | 16 (10, 22) | 20 (12, 27) | <0.001 |
| **Total Valproate, mg/L** | 46 (34, 63) | 45 (34, 62) | 47 (35, 65) | 0.35 |
| **Free Fraction** | 35% (25, 53) | 33% (23, 51) | 40% (28, 53) | 0.004 |
| **Valproate Administered in Prior 24 Hours, mg** | 1,600 (1,000, 2,500) | 1,750 (1,250, 2,500) | 1,500 (1,000, 2,500) | 0.020 |
| **Albumin, g/dL** | 2.80 (2.50, 3.10) | 2.80 (2.50, 3.20) | 2.80 (2.50, 3.00) | 0.75 |
| Unknown | 45 | 36 | 9 | |
| **BUN, mg/dL** | 24 (16, 40) | 25 (16, 42) | 20 (13, 30) | <0.001 |
| Unknown | 3 | 2 | 1 | |
| **SCr, mg/dL** | 0.81 (0.57, 1.47) | 0.86 (0.61, 1.81) | 0.68 (0.51, 1.08) | <0.001 |
| Unknown | 3 | 2 | 1 | |
| **Meds Within 24 Hours of Concentration Measurement** | | | | |
| Propofol | 159 (29%) | 105 (27%) | 54 (34%) | 0.073 |
| Phenytoin | 11 (2.0%) | 4 (1.0%) | 7 (4.5%) | 0.015 |
| Carbapenem | 17 (3.1%) | 13 (3.3%) | 4 (2.5%) | 0.79 |
| Clevidipine | 4 (0.7%) | 3 (0.8%) | 1 (0.6%) | >0.99 |
| Aspirin | 122 (22%) | 89 (23%) | 33 (21%) | 0.68 |
| Acetaminophen | 279 (51%) | 205 (52%) | 74 (47%) | 0.29 |
| Ibuprofen | 4 (0.7%) | 4 (1.0%) | 0 (0%) | 0.58 |
| Ketorolac | 1 (0.2%) | 0 (0%) | 1 (0.6%) | 0.29 |
| **Therapeutically Concordant** | 88 (16%) | 67 (17%) | 21 (13%) | 0.29 |

[1]Median (IQR); n (%)

[2]Wilcoxon rank sum test; Pearson's Chi-squared test; Fisher's exact test

BUN: blood urea nitrogen

**Figure S1: Protein Binding Percentage versus Total Valproate Concentration**

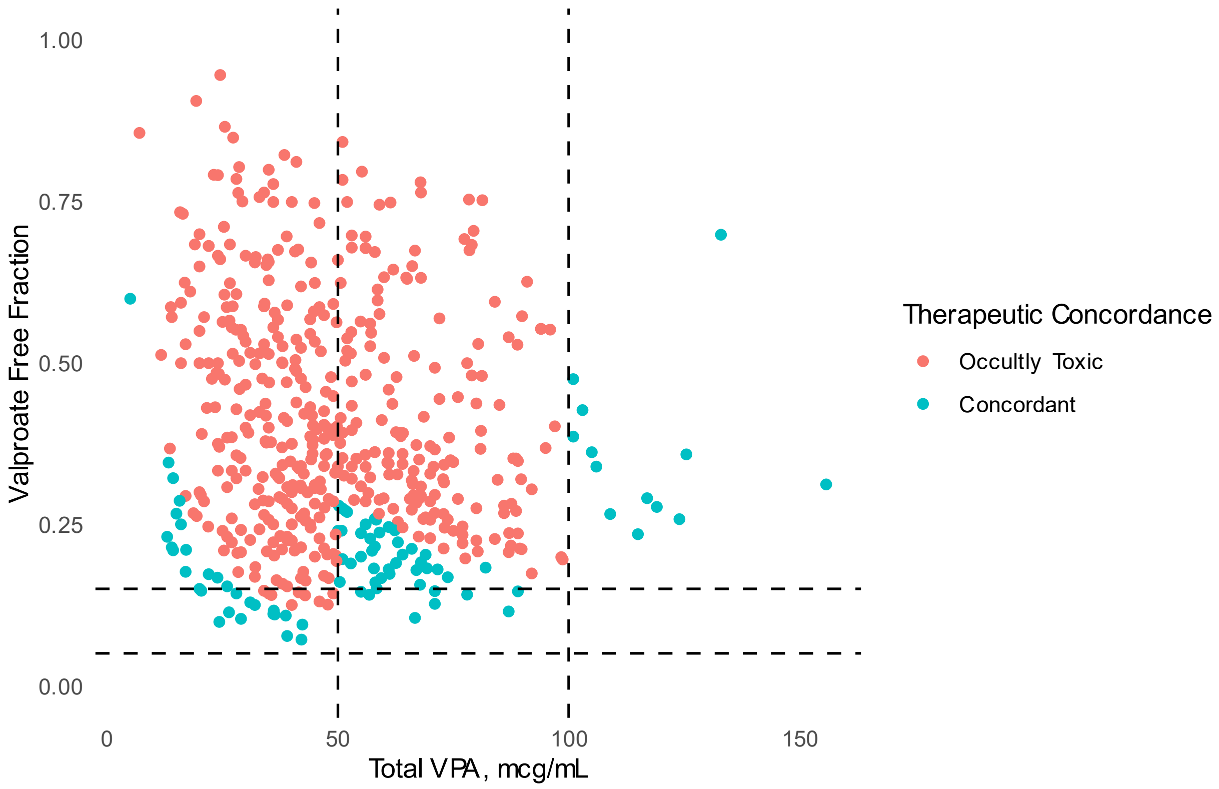


*Vertical lines represent therapeutic range of total valproate concentrations (50-100 mcg/mL) and horizontal lines represent normal fraction of unbound valproate (5-15%)*

**Figure S2: Relationship Between Highest Free Valproate Concentration and Lowest Platelet Count**

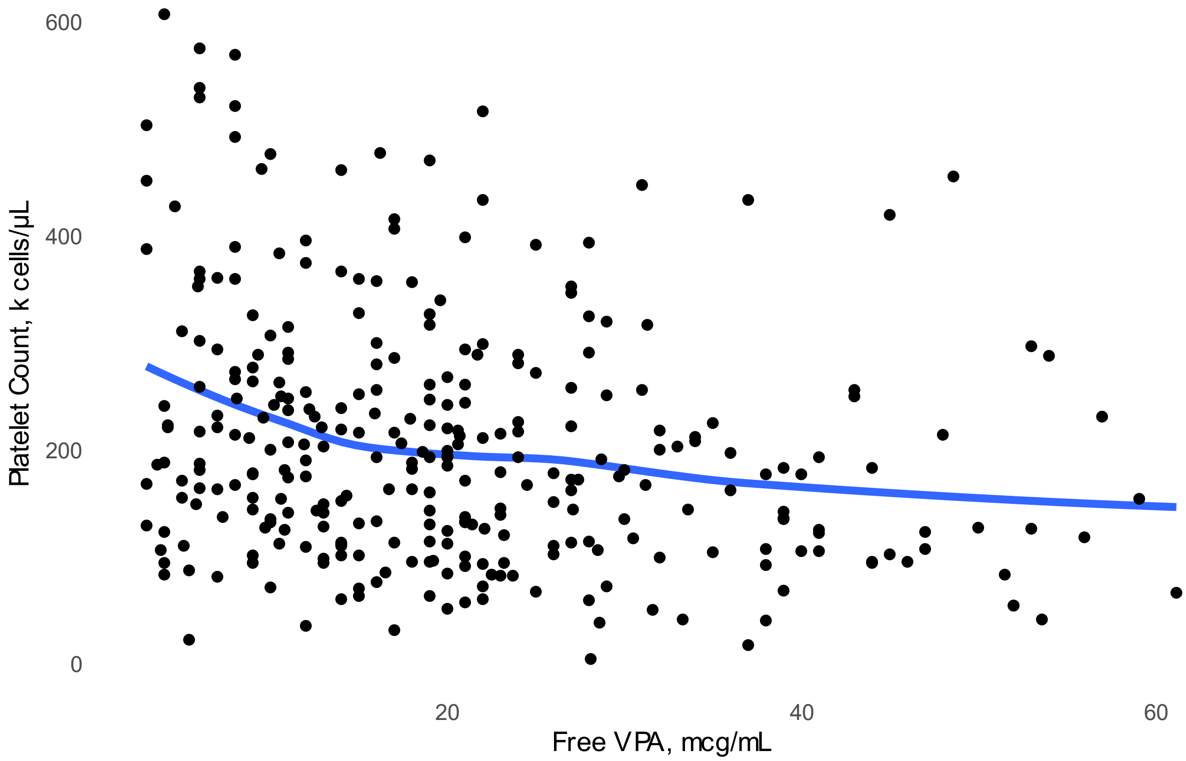


*The lowest platelet count measured within seven days of the maximum free valproate concentration were included. Trend line represents a loess regression line. One outlier (free valproate concentration of 93 mcg/mL) was removed from the chart but did not significantly impact the shape or slope of the line (associated platelet count of 163 x 10 $^{9}$).*

**Figure S3: Performance of Serum Albumin as a Determinant for Therapeutic Discordance**

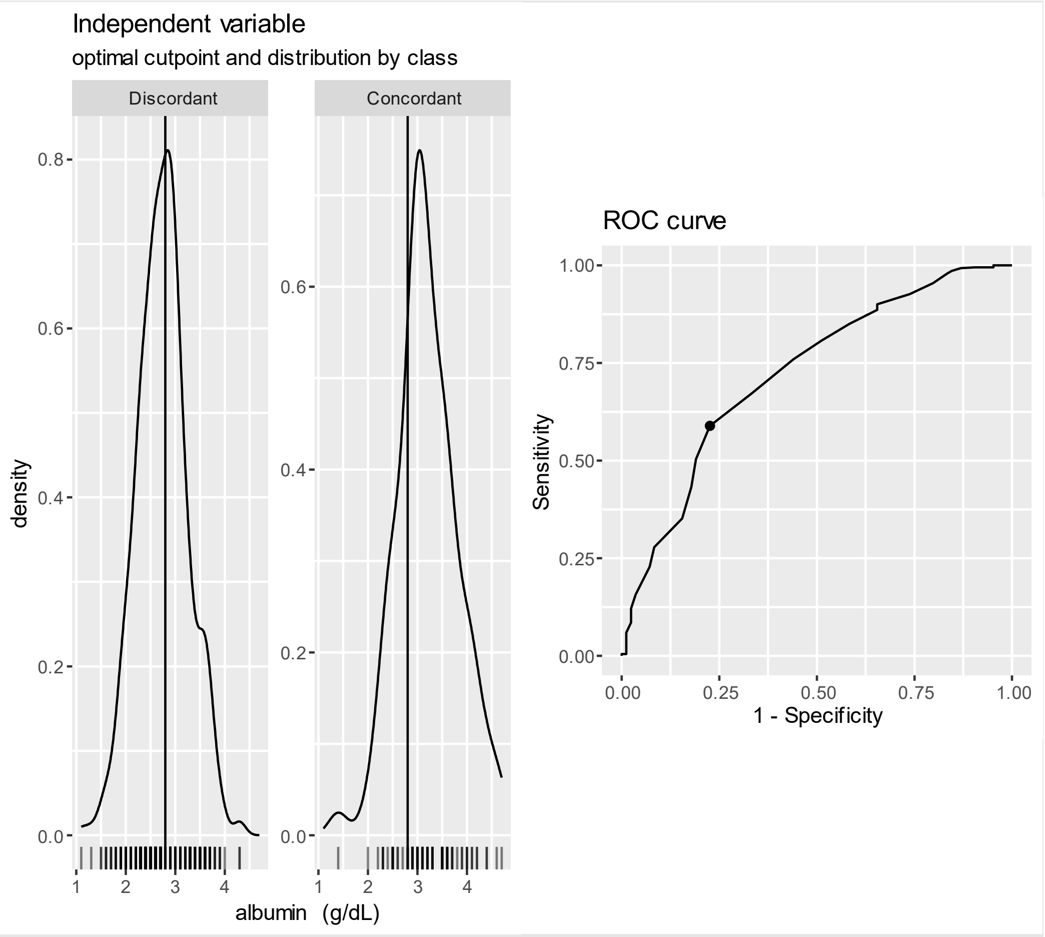


*Albumin included in a univariate logistic regression model for occult toxicity. The Receive-Operator Characteristic (ROC) curve represents the sensitivity and specificity of serum albumin for predicting occult toxicity, with an optimal cutpoint of 2.9 g/dL identified as the best tradeoff between sensitivity and specificity. An albumin of less than 2.9 g/dL was associated with a 92.8% "positive" predictive value for discordance and a 27.3% "negative" predictive value for concordance.*

### Figure S4: Performance of Free Valproate as a Determinant for Thrombocytopenia

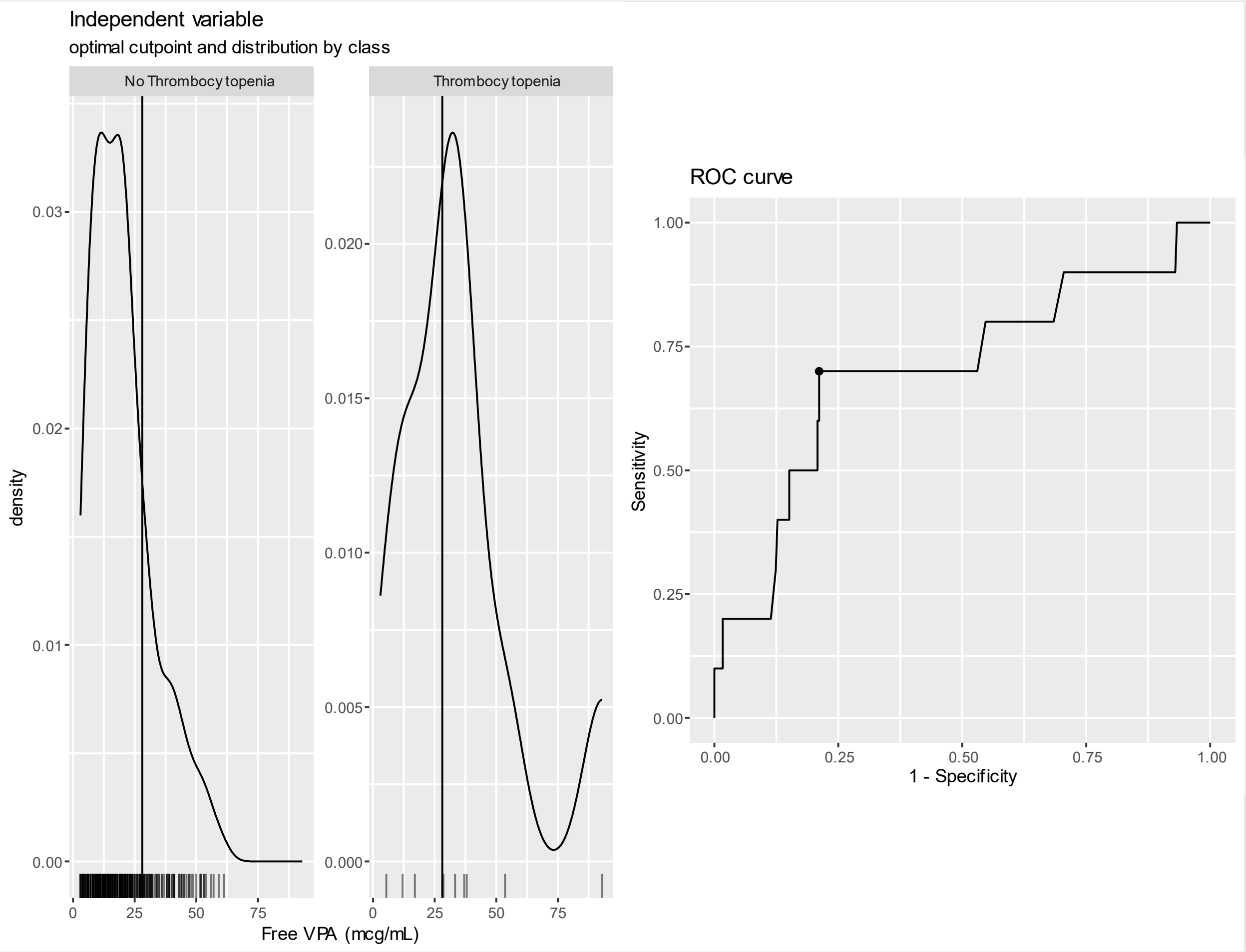


*Free valproate included in a univariate logistic regression model for thrombocytopenia (platelet count <50,000). The Receive-Operator Characteristic (ROC) curve represents the sensitivity and specificity of free valproate for predicting thrombocytopenia, with an optimal cutpoint of 28.1 mcg/mL identified as the best tradeoff between sensitivity and specificity (AUC 0.70). A free valproate of greater than 28.1 mcg/mL was associated with a 10% positive predictive value for thrombocytopenia and concentrations less than 28.1 mcg/mL were associated with a 98.7% negative predictive value for thrombocytopenia.*

### Figure S4S5: Performance of Free Valproate as a Determinant for Hepatotoxicity

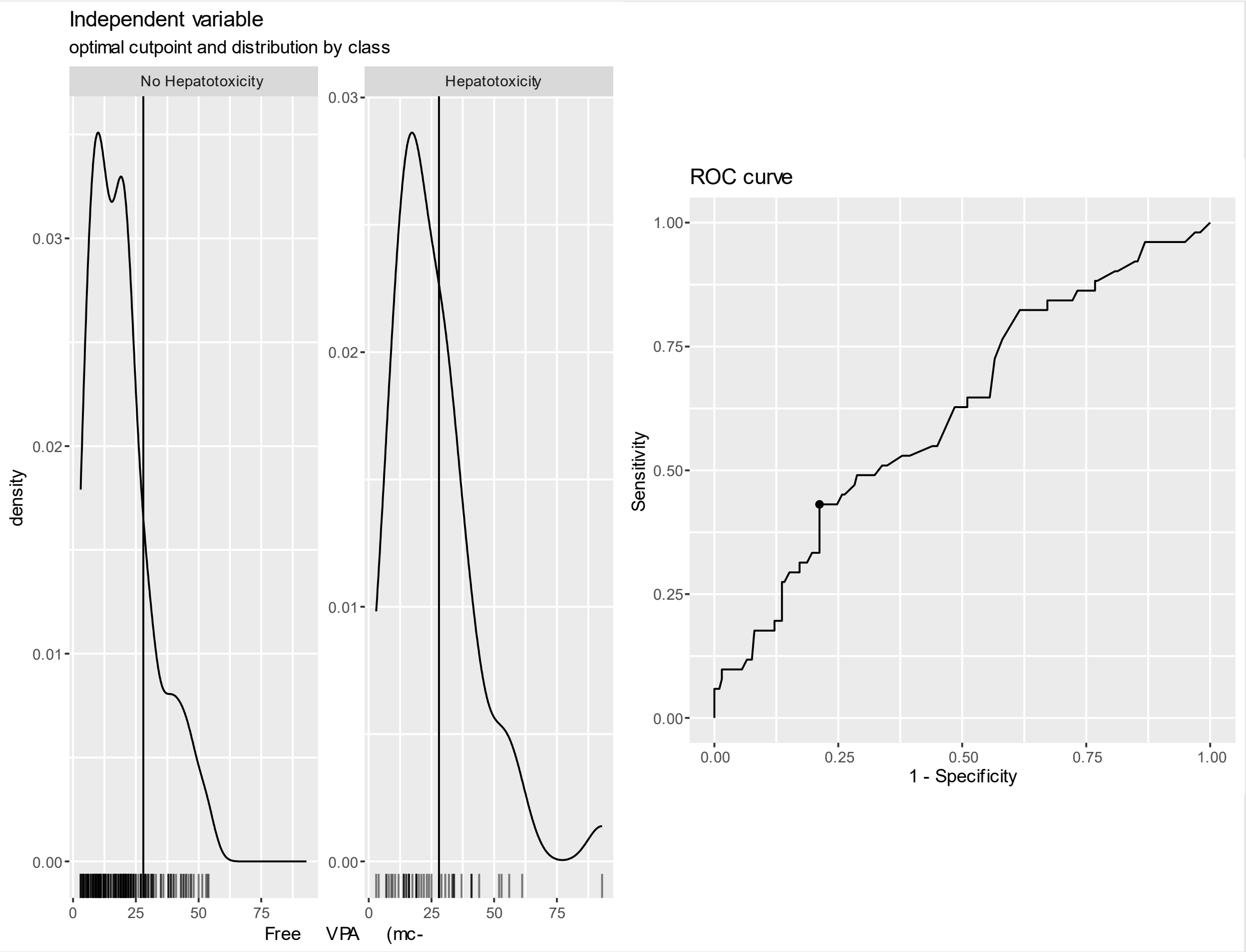


*Free valproate included in a univariate logistic regression model for hepatotoxicity. The Receive-Operator Characteristic (ROC) curve represents the sensitivity and specificity of free valproate for predicting thrombocytopenia, with an optimal cutpoint of 28 mcg/mL identified as the best tradeoff between sensitivity and specificity (AUC 0.62). A free valproate of greater than 28 mcg/mL was associated with a 34% positive predictive value for hepatotoxicity and concentrations less than 28 mcg/mL were associated with an 84.3% negative predictive value for hepatotoxicity.*
